# The effectiveness of malaria camps as part of the malaria control program in Odisha, India

**DOI:** 10.1101/2023.01.18.23284743

**Authors:** Danielle C. Ompad, Timir K. Padhan, Anne Kessler, Stuti Mohanty, Yesim Tozan, Abbey M. Jones, Anna Maria van Eijk, Steven A. Sullivan, Mohammed A. Haque, Madan Mohan Pradhan, Sanjib Mohanty, Jane M. Carlton, Praveen K. Sahu

**Author notes:** Correspondence and requests for materials should be addressed to Danielle Ompad and Praveen Sahu. TKP and AK are joint second authors. PKS, JMC, SM, and MMP are joint last authors.

## Abstract

Durgama Anchalare Malaria Nirakaran (DAMaN) is a multi-component malaria intervention for hard-to-reach villages in Odisha, India. The main component, Malaria Camps (MCs), consists of mass screening, treatment, education, and intensified vector control. We evaluated MC effectiveness using a quasi-experimental cluster-assigned stepped-wedge study with a pretest-posttest control group in 15 villages: six immediate (Arm A), six delayed (Arm B), and three previous interventions (Arm C). The primary outcome was PCR+ *Plasmodium* infection prevalence. Across all arms, the odds of PCR+ malaria were 54% lower at the third follow-up compared to baseline. A time (i.e., visit) x study arm interaction revealed significantly lower odds of PCR+ malaria in Arm A versus B at the third follow-up. The cost per person ranged between US$3-8, the cost per tested US$4-7, and the cost per treated US$82-1,614, per camp round. These results suggest that the DAMaN intervention is a promising, financially feasible approach for malaria control.

## Introduction

India has made noteworthy progress towards malaria elimination, with cases decreasing from 20 million in 2000 to approximately 4.1 million cases in 2020^1^. Despite this decline over the past two decades, it remains an important public health problem. India was one of eleven countries accounting for 70% of the global burden of malaria in 2020^1^, and it continues to account for 79% of all malaria cases and 83% of all malaria deaths in the South-East Asia region^2^. Within India, the state of Odisha has the highest burden of malaria, accounting for 22.4% of all cases in the country in 2020, of which 91.4% were *Plasmodium falciparum* infections^3^. The malaria burden in Odisha has been persistently high in the remote, forested areas of the state. In response to this, the Government of Odisha implemented the Durgama Anchalare Malaria Nirakaran (DAMaN; ‘malaria control in inaccessible areas’) program in 2017. DAMaN was designed to supplement existing and routine malaria control programs that serve approximately 5000 inaccessible villages and hamlets in the rural parts of the state^4^.

A key activity of the DAMaN program is the implementation of ‘malaria camps’ (MCs). In the MC model, teams of health workers visit villages and hamlets to provide the intervention which includes seven key activities in each cycle: (1) one round of mass screen and treat (MSAT) before the monsoon season conducted with point-of-care rapid diagnostic tests (RDTs), (2) one or two rounds of fever screen and treat (FSAT) during or after the monsoon season, (3) IRS (indoor residual spraying), (4) other vector control methods, (5) LLIN (long-lasting insecticidal net) distribution (not on an annual basis), (6) educational programming, and (7) maternal and child health visits and screenings. The intervention has been described in detail^4, 5, 6, 7^. The control condition, as a part of the National Vector Control Strategy, is standard of care (SOC) whereby Accredited Social Health Activists (ASHAs) conduct door-to-door fever surveillance weekly. Fever cases are tested with RDTs and treated; severe cases are referred to hospitals^8^. IRS is conducted twice in a year during the transmission season in selected high risk areas having an annual parasite index (API) >5, and LLINs are distributed in high risk areas having API>2^9^. Thus key distinguishing features of MCs versus SOC include MSAT (everyone is offered malaria screening regardless of fever status in the MCs), educational programming, and maternal and child health visits and screenings.

Odisha state has seen a >80% decline in malaria cases since 2017, attributed to the large-scale distribution of LLINs and the DAMaN program^10^. The aim of our project was to evaluate the effectiveness of the DAMaN MCs through a quasi-experimental cluster-assigned stepped-wedge study with a pretest-posttest control group design. We enrolled 2463 participants into three study arms: six immediate interventions (Arm A), six delayed interventions (Arm B), and three previous interventions (Arm C), and sampled them at baseline, with three follow-ups from August 2019 to December 2020. The primary outcome was PCR+ *Plasmodium* infection prevalence. Across all arms, the odds of PCR+ malaria were 54% lower at the third follow-up compared to baseline. A time (i.e., visit) x study arm interaction revealed significantly lower odds of PCR+ malaria in Arm A versus B at the third follow-up. Thus our results suggest that the DAMaN program’s malaria camps were associated with lower malaria incidence relative to standard-of-care.

## Results

### Study population and study design

Fifteen villages were selected in the northern districts of Keonjhar and Jharsuguda in Odisha state, India, in consultation with the Odisha Malaria Control Program (**Figure 1**). The villages were distributed between three study arms: six villages in Arm A ‘new-MC’ (communities receiving MCs for the first time in year one), six villages in Arm B ‘delayed-MC’ (communities undergoing routine malaria control in year one and receiving MCs for the first time in year two), and three villages in Arm C ‘old-MC’ villages, where MCs had already been implemented prior to the study period. A flow chart of the study is shown in **Figure 2**.

**Figure 1.**
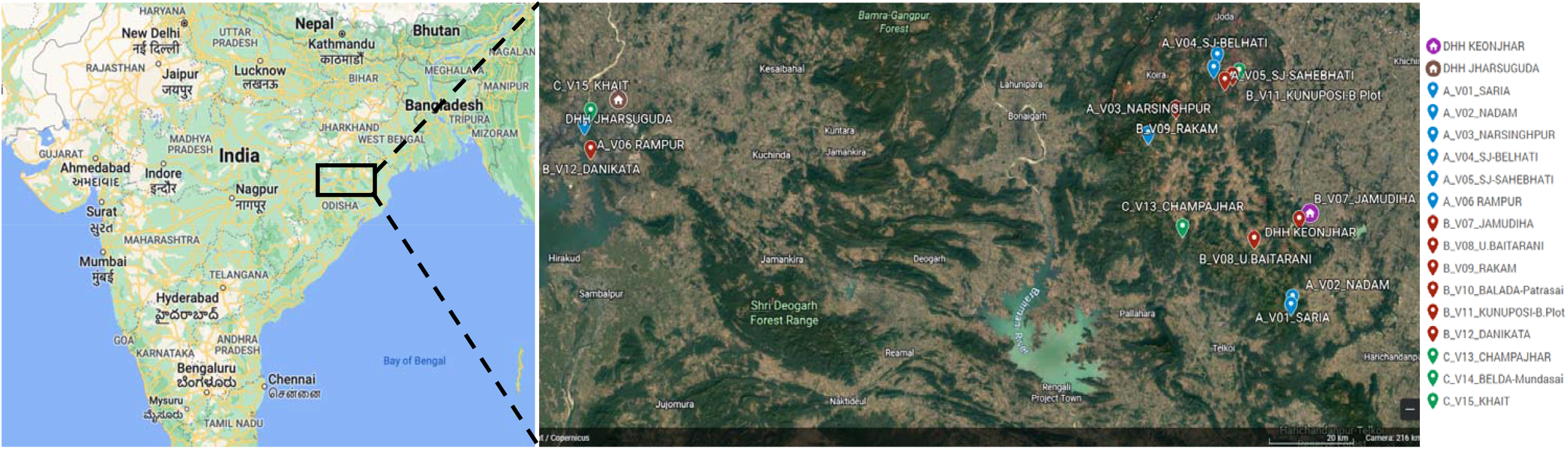
Location of 15 hard-to-reach villages in Odisha, India. The two DHH (district headquarter hospitals) are indicated, and each village is named with its study arm letter (A, B, C) and village number (V1-15) provided. Maps created with Google©2023, INEGI.

**Figure 2.**
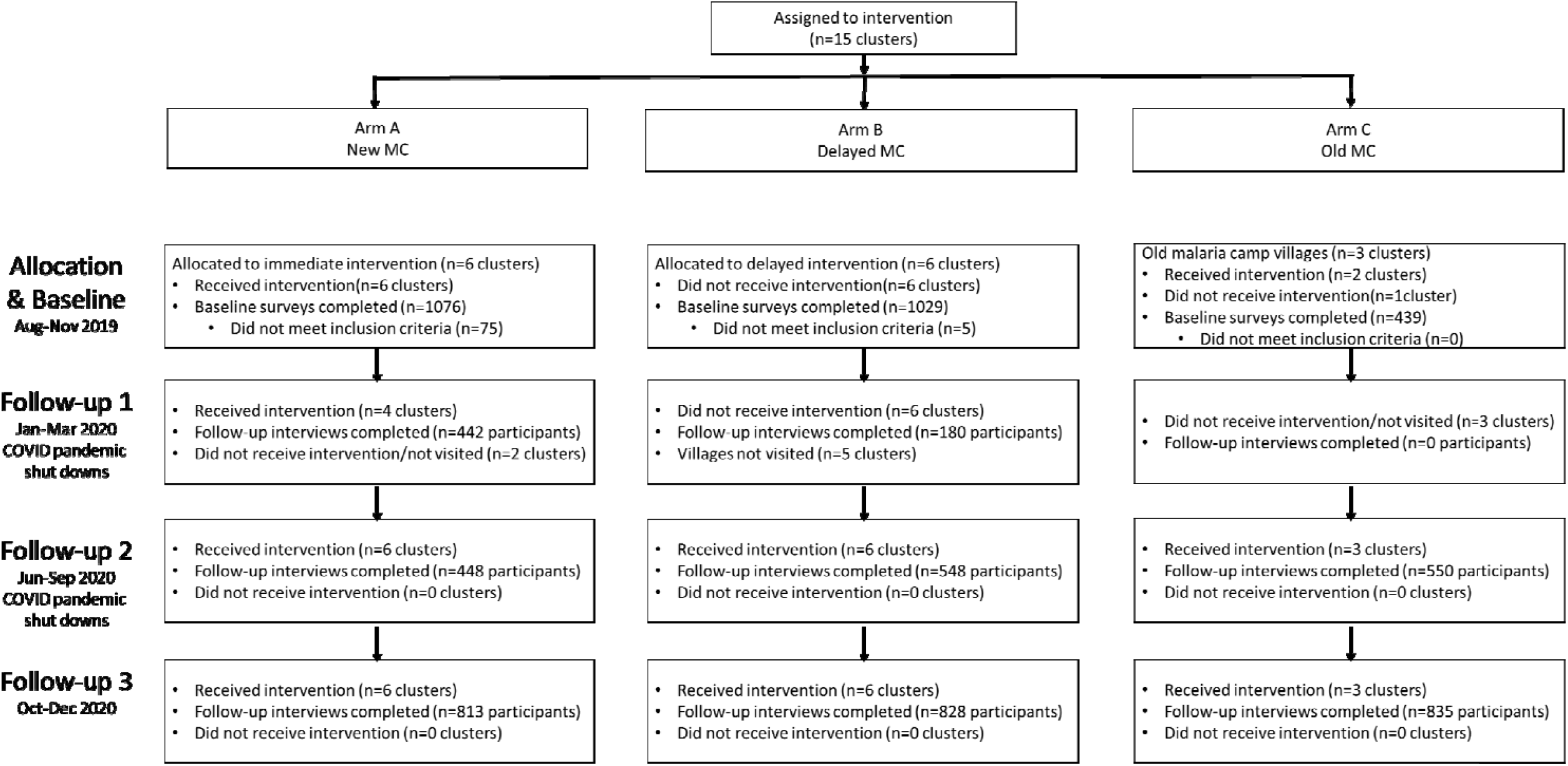
Cluster-assigned quasi-experimental study of malaria camps as part of the DAMaN malaria elimination program in Odisha, India. Flow chart providing a summary of the 15 clusters (villages) distributed between three arms, and sampled at baseline and three follow-ups. A timeline of the activities is also given, and whether COVID-19 shutdowns are known to have occurred.

We enrolled 2463 participants in the study and **Table 1** presents their baseline demographic characteristics. A total of 14% of the participants was aged 5 years or younger, 29.4% were aged 6 to 17, 23.6% were 18-35, and 32.9% were over age 35, with females comprising 56.1% of the participants. The participants generally had low educational attainment: almost half (49.4%) had no schooling/less than primary school whereas 5.6% had higher secondary school or more. With respect to occupation, 28.8% were housewives, 34.9% were students, 5.6% were farmers/agricultural laborers, 18.5% were employed in another trade, 9.6% were children that were not in school, and 2.6% did not have an occupation.

**Table 1.**
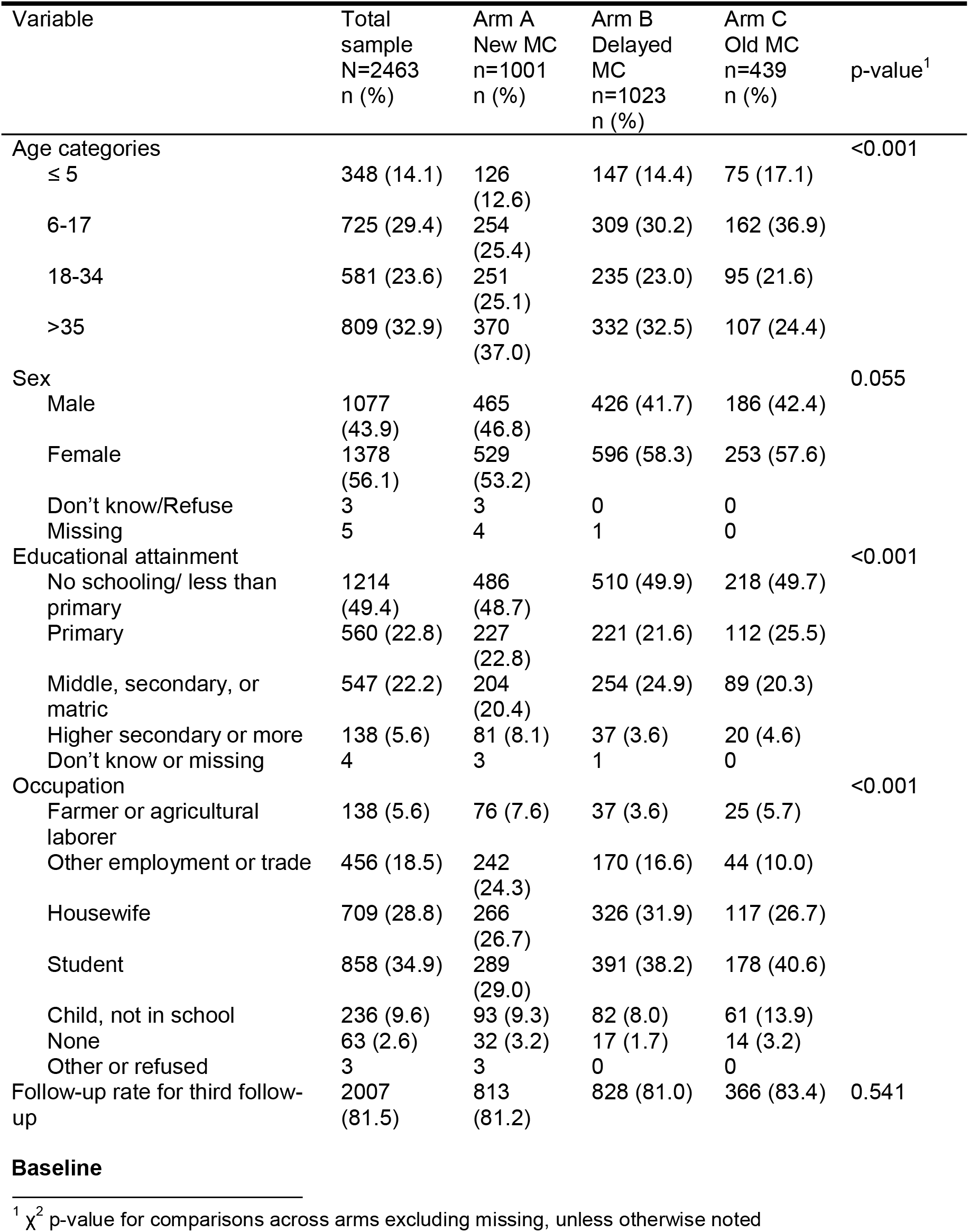
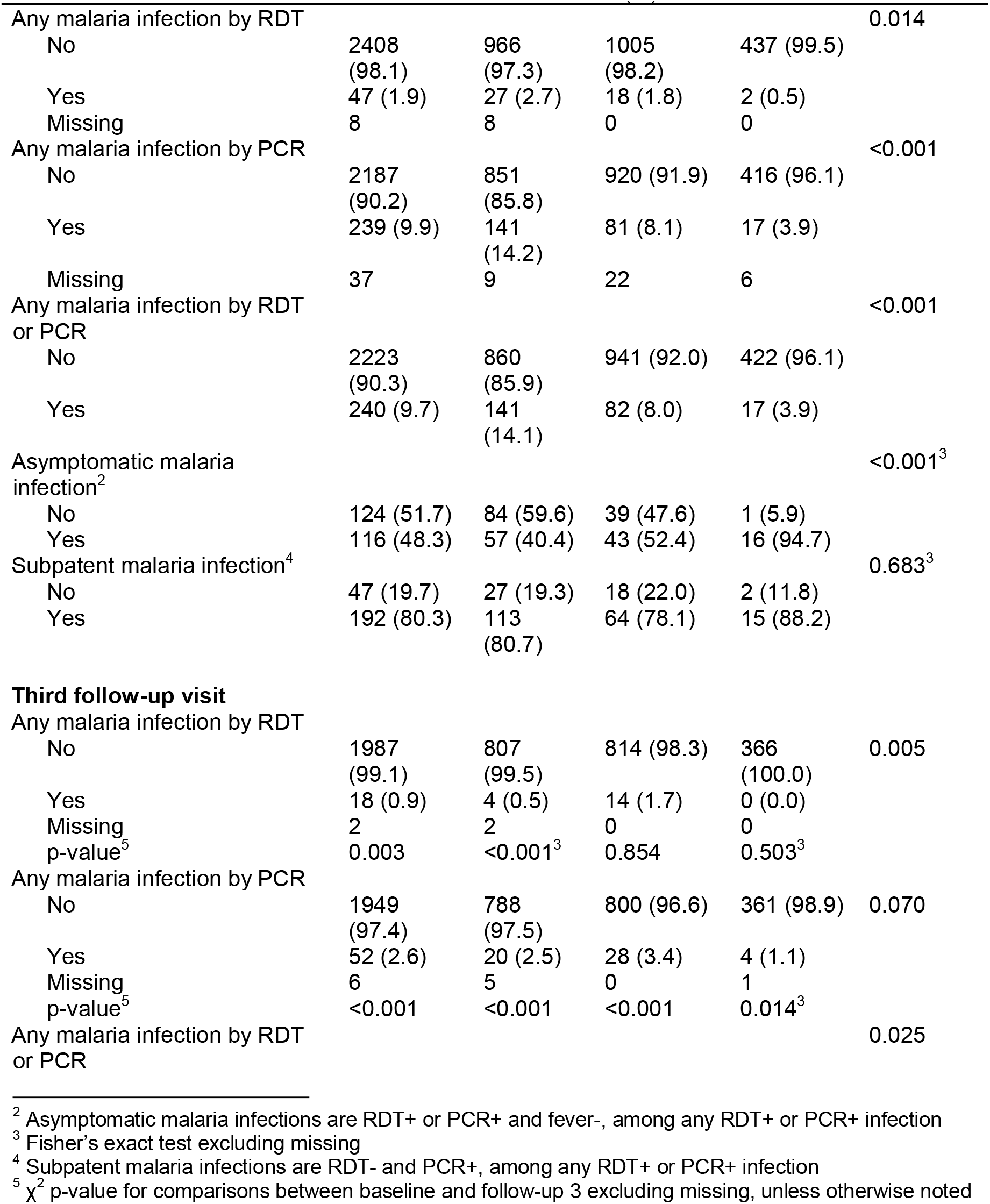
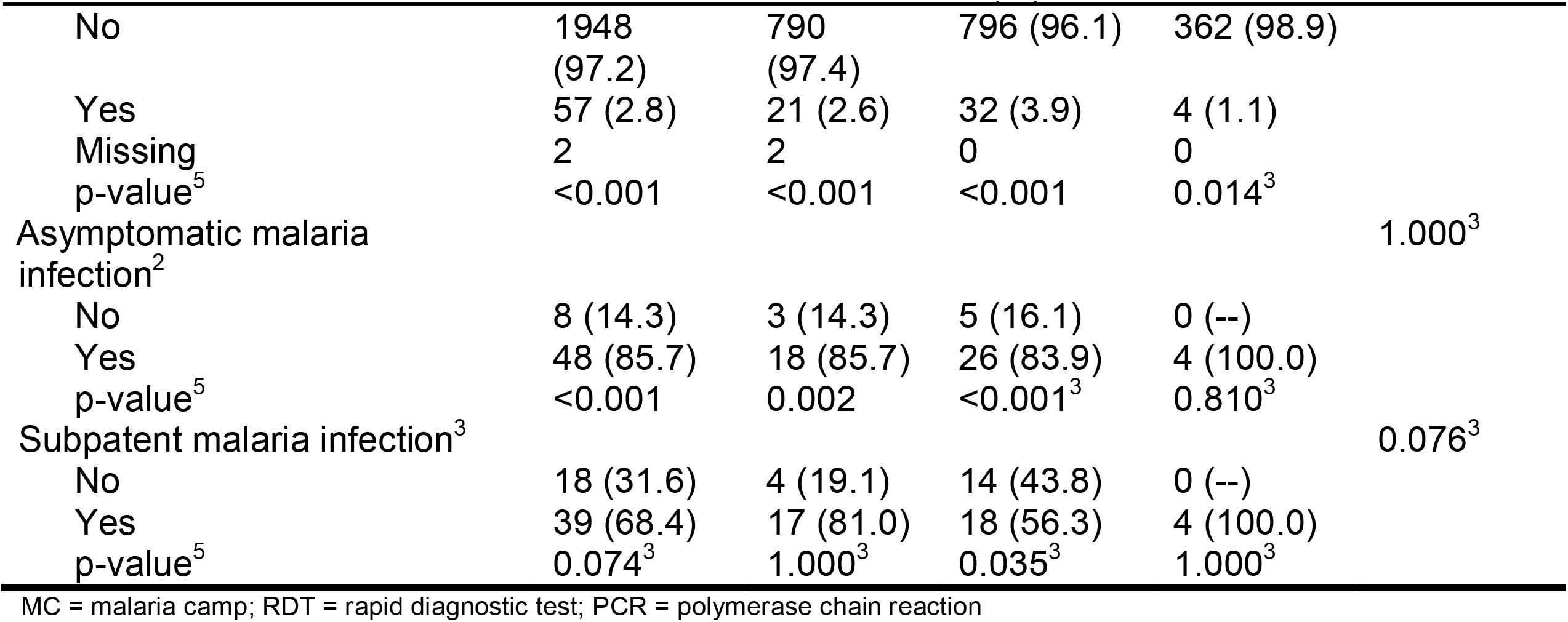
Demographic characteristics and malaria outcomes at baseline and the third follow-up visit.

| Variable | Total sample<br>N=2463<br>n (%) | Arm A<br>New MC<br>n=1001<br>n (%) | Arm B<br>Delayed<br>MC<br>n=1023<br>n (%) | Arm C<br>Old MC<br>n=439<br>n (%) | p-value<sup>1</sup> |
| --- | --- | --- | --- | --- | --- |
| Age categories |  |  |  |  | <0.001 |
| ≤ 5 | 348 (14.1) | 126<br>(12.6) | 147 (14.4) | 75 (17.1) |  |
| 6-17 | 725 (29.4) | 254<br>(25.4) | 309 (30.2) | 162 (36.9) |  |
| 18-34 | 581 (23.6) | 251<br>(25.1) | 235 (23.0) | 95 (21.6) |  |
| >35 | 809 (32.9) | 370<br>(37.0) | 332 (32.5) | 107 (24.4) |  |
| Sex |  |  |  |  | 0.055 |
| Male | 1077<br>(43.9) | 465<br>(46.8) | 426 (41.7) | 186 (42.4) |  |
| Female | 1378<br>(56.1) | 529<br>(53.2) | 596 (58.3) | 253 (57.6) |  |
| Don't know/Refuse | 3 | 3 | 0 | 0 |  |
| Missing | 5 | 4 | 1 | 0 |  |
| Educational attainment |  |  |  |  | <0.001 |
| No schooling/ less than<br>primary | 1214<br>(49.4) | 486<br>(48.7) | 510 (49.9) | 218 (49.7) |  |
| Primary | 560 (22.8) | 227<br>(22.8) | 221 (21.6) | 112 (25.5) |  |
| Middle, secondary, or<br>matric | 547 (22.2) | 204<br>(20.4) | 254 (24.9) | 89 (20.3) |  |
| Higher secondary or more | 138 (5.6) | 81 (8.1) | 37 (3.6) | 20 (4.6) |  |
| Don't know or missing | 4 | 3 | 1 | 0 |  |
| Occupation |  |  |  |  | <0.001 |
| Farmer or agricultural<br>laborer | 138 (5.6) | 76 (7.6) | 37 (3.6) | 25 (5.7) |  |
| Other employment or trade | 456 (18.5) | 242<br>(24.3) | 170 (16.6) | 44 (10.0) |  |
| Housewife | 709 (28.8) | 266<br>(26.7) | 326 (31.9) | 117 (26.7) |  |
| Student | 858 (34.9) | 289<br>(29.0) | 391 (38.2) | 178 (40.6) |  |
| Child, not in school | 236 (9.6) | 93 (9.3) | 82 (8.0) | 61 (13.9) |  |
| None | 63 (2.6) | 32 (3.2) | 17 (1.7) | 14 (3.2) |  |
| Other or refused | 3 | 3 | 0 | 0 |  |
| Follow-up rate for third follow-up | 2007<br>(81.5) | 813<br>(81.2) | 828 (81.0) | 366 (83.4) | 0.541 |
| Any malaria infection by RDT |  |  |  |  | 0.014 |
| No | 2408<br>(98.1) | 966<br>(97.3) | 1005<br>(98.2) | 437 (99.5) |  |
| Yes | 47 (1.9) | 27 (2.7) | 18 (1.8) | 2 (0.5) |  |
| Missing | 8 | 8 | 0 | 0 |  |
| Any malaria infection by PCR |  |  |  |  | <0.001 |
| No | 2187<br>(90.2) | 851<br>(85.8) | 920 (91.9) | 416 (96.1) |  |
| Yes | 239 (9.9) | 141<br>(14.2) | 81 (8.1) | 17 (3.9) |  |
| Missing | 37 | 9 | 22 | 6 |  |
| Any malaria infection by RDT<br>or PCR |  |  |  |  | <0.001 |
| No | 2223<br>(90.3) | 860<br>(85.9) | 941 (92.0) | 422 (96.1) |  |
| Yes | 240 (9.7) | 141<br>(14.1) | 82 (8.0) | 17 (3.9) |  |
| Asymptomatic malaria<br>infection<sup>2</sup> |  |  |  |  | <0.001<sup>3</sup> |
| No | 124 (51.7) | 84 (59.6) | 39 (47.6) | 1 (5.9) |  |
| Yes | 116 (48.3) | 57 (40.4) | 43 (52.4) | 16 (94.7) |  |
| Subpatent malaria infection<sup>4</sup> |  |  |  |  | 0.683<sup>3</sup> |
| No | 47 (19.7) | 27 (19.3) | 18 (22.0) | 2 (11.8) |  |
| Yes | 192 (80.3) | 113<br>(80.7) | 64 (78.1) | 15 (88.2) |  |
| **Third follow-up visit** |  |  |  |  |  |
| Any malaria infection by RDT |  |  |  |  |  |
| No | 1987<br>(99.1) | 807<br>(99.5) | 814 (98.3) | 366<br>(100.0) | 0.005 |
| Yes | 18 (0.9) | 4 (0.5) | 14 (1.7) | 0 (0.0) |  |
| Missing | 2 | 2 | 0 | 0 |  |
| p-value<sup>5</sup> | 0.003 | <0.001<sup>3</sup> | 0.854 | 0.503<sup>3</sup> |  |
| Any malaria infection by PCR |  |  |  |  |  |
| No | 1949<br>(97.4) | 788<br>(97.5) | 800 (96.6) | 361 (98.9) | 0.070 |
| Yes | 52 (2.6) | 20 (2.5) | 28 (3.4) | 4 (1.1) |  |
| Missing | 6 | 5 | 0 | 1 |  |
| p-value<sup>5</sup> | <0.001 | <0.001 | <0.001 | 0.014<sup>3</sup> |  |
| Any malaria infection by RDT<br>or PCR |  |  |  |  | 0.025 |

<sup>2</sup> Asymptomatic malaria infections are RDT+ or PCR+ and fever-, among any RDT+ or PCR+ infection
<sup>3</sup> Fisher's exact test excluding missing
<sup>4</sup> Subpatent malaria infections are RDT- and PCR+, among any RDT+ or PCR+ infection
<sup>5</sup> $\chi^2$ p-value for comparisons between baseline and follow-up 3 excluding missing, unless otherwise noted

| Variable | Total sample<br>N=2463<br>n (%) | Arm A<br>New MC<br>n=1001<br>n (%) | Arm B<br>Delayed<br>MC<br>n=1023<br>n (%) | Arm C<br>Old MC<br>n=439<br>n (%) | p-value<sup>1</sup> |
| --- | --- | --- | --- | --- | --- |
| No | 1948<br>(97.2) | 790<br>(97.4) | 796 (96.1) | 362 (98.9) |  |
| Yes | 57 (2.8) | 21 (2.6) | 32 (3.9) | 4 (1.1) |  |
| Missing | 2 | 2 | 0 | 0 |  |
| p-value<sup>5</sup> | <0.001 | <0.001 | <0.001 | 0.014<sup>3</sup> |  |
| Asymptomatic malaria<br>infection<sup>2</sup> |  |  |  |  | 1.000<sup>3</sup> |
| No | 8 (14.3) | 3 (14.3) | 5 (16.1) | 0 (--) |  |
| Yes | 48 (85.7) | 18 (85.7) | 26 (83.9) | 4 (100.0) |  |
| p-value<sup>5</sup> | <0.001 | 0.002 | <0.001<sup>3</sup> | 0.810<sup>3</sup> |  |
| Subpatent malaria infection<sup>3</sup> |  |  |  |  | 0.076<sup>3</sup> |
| No | 18 (31.6) | 4 (19.1) | 14 (43.8) | 0 (--) |  |
| Yes | 39 (68.4) | 17 (81.0) | 18 (56.3) | 4 (100.0) |  |
| p-value<sup>5</sup> | 0.074<sup>3</sup> | 1.000<sup>3</sup> | 0.035<sup>3</sup> | 1.000<sup>3</sup> |  |

MC = malaria camp; RDT = rapid diagnostic test; PCR = polymerase chain reaction

Across the study arms there were significant differences in age, educational attainment, and occupation, but not sex. As evidenced by the age category distribution and the mean age for each arm, Arm A was older (mean age [M_age_]=27.2, standard deviation [SD]=19.0) and Arm C was younger (M_age_=22.2, SD=18.8) than Arm B (M_age_ = 25.7, SD = 19.6, p_ANOVA_<0.001). A higher proportion of participants in Arm A had higher secondary education or higher while a higher proportion in Arm B had middle, secondary, or matric and in Arm C a high proportion had primary education. There were a higher proportion of farmers/agricultural laborers and people with other employment/trades in Arm A, a higher proportion of housewives in Arm B, and higher proportions of students and children not in school in Arm C. Given the differences across arms, adjusted odds ratios (aORs) control for demographics.

### Intent-to-treat analysis for primary and secondary malaria outcomes

The MC intervention was revised from our initial protocol paper^7^ with two major and consistent differences. First, all MCs conducted MSAT rather than FSAT in all rounds of the intervention. Second, LLINs were not distributed in our study villages because their 3-yearly replenishment distribution scheduled for 2020 was delayed due to the Covid-19 pandemic.

**Table 1** presents the malaria outcomes at baseline and the third follow-up visit; associations are considered significant if p<0.013. At baseline, 9.9% of the sample had PCR+ *Plasmodium* infection, with significant differences across study arms. Arm A had the highest prevalence of *Plasmodium* infection (14.2%), followed by Arm B (8.1%) and Arm C (3.9%, p<0.001). Prevalence of RDT+ *Plasmodium* infection was 1.9% and there were no significant differences across arms; Arm A again had the highest prevalence of *Plasmodium* infection (2.7%), followed by Arm B (1.8%) and Arm C (0.5%, p=0.014). When malaria was defined by any positive PCR or RDT test, prevalence overall was 9.9% at baseline. With respect to species, 236 (98.3%) were *P. falciparum* cases and 9 (3.8%) were *P. vivax*; 5 (2.1%) were co-infected with *P. falciparum* and *P. vivax* [data not shown]. Approximately 48.3% of malaria infections were asymptomatic (defined as RDT+ or PCR+ and no fever) at baseline with Arm C having the highest proportion (94.7%) of asymptomatic cases as compared to Arms A and B (40.4 and 52.4%, respectively; p<0.001). A substantial majority (80.3%) of malaria cases were subpatent (i.e., RDT- and PCR+), with no significant differences across arms.

At the third follow-up visit, 2.6% of the sample had PCR+ *Plasmodium* infection, with no significant differences across arms. Prevalence of RDT+ *Plasmodium* infection was 0.9% overall at follow-up, with 0.5% prevalence in Arm A, 1.7% prevalence in Arm B, and no observed cases in Arm C (p=0.005). When malaria was defined by any positive PCR or RDT test, prevalence overall was 2.8% at baseline. When we compared malaria outcomes from baseline to follow-up for the entire sample as well as each arm, we found that there was a significant reduction in both RDT+ and PCR+ *Plasmodium* infection in the sample overall and for Arm A. We observed a significant reduction in PCR+ but not RDT+ *Plasmodium* infection in Arm B and no significant differences in Arm C. With respect to species, 236 (98.3%) were *P. falciparum* cases and 9 (3.8%) were *P. vivax*; 5 (2.1%) were co-infected with *P. falciparum* and *P. vivax* [data not shown]. Approximately 85.7% of malaria infections were asymptomatic at FU3 with no significant differences across arms but significant increases in the proportion of cases that were asymptomatic from baseline to FU3 for the entire sample (p<0.001), Arm A (p=0.002), and Arm B (p<0.001) but not Arm C. A substantial majority (68.4%) of malaria cases were subpatent at FU3, with a significant decrease from baseline to FU3 in Arm B only (78.1 to 56.3%, p=0.035).

The overall follow-up rate for FU3 was 81.5% with a range of 81.5 to 83.4% across arms, with no significant differences across arms (**Table 1**). The sociodemographic and malaria outcome correlates of participants lost-to-follow-up is shown in **Table 2**.

There were no significant differences in loss-to-follow-up (LTFU) by study arm or sex. Younger people were more likely to be LTFU as were those with lower educational attainment, students, and children not in school. Those with malaria, regardless of detection method, were significantly more likely to be LTFU but there were no significant differences by asymptomatic or subpatent cases.

**Table 2.** Demographic characteristics and malaria outcomes for individuals retained in the study versus those lost-to-follow-up.

| Variable | Lost-to-follow-up |  | p-value<sup>1</sup> |
| --- | --- | --- | --- |
|  | No<br>n=2,007<br>n (%) | Yes<br>n=456<br>n (%) |  |
| Study arm |  |  |  |
| Arm A (New MC) | 813 (40.5) | 188 (41.2) | 0.526 |
| Arm B (Delayed MC) | 828 (41.3) | 195 (42.8) |  |
| Arm C (Old MC) | 366 (18.2) | 73 (16.0) |  |
| Age categories |  |  |  |
| ≤5 | 252 (12.6) | 96 (21.1) | <0.001 |
| 6-17 | 571 (28.5) | 154 (33.8) |  |
| 18-34 | 500 (24.9) | 81 (17.8) |  |
| ≥35 | 684 (34.1) | 125 (27.4) |  |
| Sex |  |  |  |
| Male | 859 (42.8) | 218 (47.8) | 0.127<sup>2</sup> |
| Female | 1141 (56.9) | 237 (51.0) |  |
| Don't know/Refuse | 2 (0.1) | 1 (0.2) |  |
| Missing | 5 (0.3) | 0 (0.0) |  |
| Educational attainment |  |  |  |
| No schooling/ less than primary | 973 (48.5) | 241 (52.9) | 0.002<sup>2</sup> |
| Primary | 453 (22.6) | 107 (23.5) |  |
| Middle, secondary, or matric | 447 (22.3) | 100 (21.9) |  |
| Higher secondary or more | 130 (6.5) | 8 (1.8) |  |
| Don't know or missing | 4 (0.2) | 0 (0.0) |  |
| Occupation |  |  |  |
| Farmer or agricultural laborer | 105 (5.2) | 33 (7.2) | <0.001 |
| Other employment or trade | 385 (19.2) | 71 (15.6) |  |
| Housewife | 616 (30.7) | 93 (20.4) |  |
| Student | 686 (34.2) | 172 (37.7) |  |
| Child, not in school | 163 (8.1) | 73 (16.0) |  |
| None | 50 (2.5) | 13 (2.9) |  |
| Other or refused | 2 (0.1) | 1 (0.2) |  |
| Any malaria by RDT |  |  |  |
| No | 1,988 (99.1) | 417 (91.5) | <0.001<sup>2</sup> |
| Yes | 12 (0.6) | 36 (7.9) |  |
| Missing | 7 (0.4) | 3 (0.7) |  |
| Any malaria by PCR |  |  |  |
| No | 1,931 (96.2) | 259 (56.8) | <0.001 |
| Yes | 51 (2.5) | 188 (41.2) |  |
| Missing | 25 (1.3) | 9 (2.0) |  |
| Any malaria by RDT or PCR |  |  |  |
| No | 1,955 (97.4) | 266 (58.3) | <0.001 |
| Yes | 52 (2.6) | 190 (41.7) |  |
| Asymptomatic malaria infection<sup>3</sup> |  |  | 0.142 |
| No | 31 (60.8) | 93 (49.2) |  |
| Yes | 20 (39.2) | 96 (50.8) |  |
| Subpatent malaria infection<sup>4</sup> |  |  | 0.752 |
| No | 11 (21.6) | 37 (19.6) |  |
| Yes | 40 (78.4) | 152 (80.4) |  |

MC = malaria camp; RDT = rapid diagnostic test; PCR = polymerase chain reaction
<sup>1</sup> $\chi^2$ p-value, unless otherwise noted
<sup>2</sup> Fishers exact test
<sup>3</sup> Asymptomatic malaria infections are RDT+ or PCR+ and fever-
<sup>4</sup> Subpatent malaria infections are RDT- and PCR+

**Table 3** presents the crude and adjusted multilevel mixed effects logistic regression models for the primary and secondary malaria outcomes. The adjusted models controlled for age and gender given baseline differences in demographics across study arms. Education and occupation were not included in the model, although they were significant in the bivariate analyses, due to multicollinearity. For the primary outcome of PCR-positive *Plasmodium* infection, the adjusted model shows almost five-fold higher odds for PCR+ *Plasmodium* infection at baseline in Arm A (new MC, adjusted OR [AOR]=5.96, 95% confidence interval [CI] = 1.04, 23.59) but not Arm C (old MC) as compared to Arm B (delayed MC). Across all three arms, the odds of PCR+ *Plasmodium* infection were 54% lower at follow-up compared to baseline (95% CI = 0.23, 0.63). The time (i.e., baseline vs. FU3) x study arm interaction term shows that there were significantly lower odds of PCR+ *Plasmodium* infection in Arm A (AOR = 0.36, 95% CI = 0.17, 0.73) but not Arm C as compared to Arm B at follow-up. The adjusted model for PCR+ or RDT+ *Plasmodium* infection showed similar results. For RDT+ *Plasmodium* infection, there were no significant differences in the odds of *Plasmodium* infection at baseline or follow-up across arms. The time x study arm interaction term shows that there were significantly lower odds of PCR+ *Plasmodium* infection in Arm A (AOR = 0.17, 95% CI = 0.04, 0.70) as compared to Arm B at follow-up. An odds ratio for Arm C at follow-up was not calculated because there were no RDT+ *Plasmodium* infections at follow-up in Arm C.

**Table 3.** Multilevel mixed-effects logistic regression models estimating the association between malaria camps and RDT+ and/or PCR+ malaria infections.

| Outcome | Malaria by RDT only |  | Malaria by PCR only |  | Malaria by RDT or PCR |  |
| --- | --- | --- | --- | --- | --- | --- |
|  | Crude OR<br>(95% CI) | Adjusted OR<br>(95% CI)<sup>1</sup> | Crude OR<br>(95% CI) | Adjusted OR<br>(95% CI)<sup>1</sup> | Crude OR<br>(95% CI) | Adjusted OR<br>(95% CI)<sup>1</sup> |
| Malaria at baseline by study arm |  |  |  |  |  |  |
| Arm B Delayed MC | 1.0 | 1.0 | 1.0 | 1.0 | 1.0 | 1.0 |
| Arm A New MC | 2.19<br>(0.15, 32.47) | 2.27<br>(0.18, 29.35) | 4.81<br>(0.98, 23.76) | 4.96<br>(1.04, 23.58) | 4.49<br>(0.96, 20.98) | 4.64<br>(1.03, 21.00) |
| Arm C Old MC | 0.34<br>(0.01, 15.32) | 0.36<br>(0.01, 12.56) | 0.49<br>(0.06, 3.72) | 0.46<br>(0.06, 3.37) | 0.48<br>(0.07, 3.48) | 0.46<br>(0.07, 3.19) |
| Malaria by Follow-up |  |  |  |  |  |  |
| Baseline | 1.0 | 1.0 | 1.0 | 1.0 | 1.0 | 1.0 |
| Follow-up 3 | 1.10<br>(0.45, 2.67) | 1.10<br>(0.47, 2.60) | 0.39<br>(0.23, 0.64) | 0.38<br>(0.23, 0.63) | 0.46<br>(0.28, 0.74) | 0.45<br>(0.28, 0.72) |
| Malaria at follow-up 3 by study arm |  |  |  |  |  |  |
| Arm B Delayed MC | 1.0 | 1.0 | 1.0 | 1.0 | 1.0 | 1.0 |
| Arm A New MC | 0.14<br>(0.03, 0.62) | 0.17<br>(0.04, 0.70) | 0.35<br>(0.17, 0.73) | 0.36<br>(0.17, 0.73) | 0.32<br>(0.16, 0.65) | 0.33<br>(0.16, 0.65) |
| Arm C Old MC | --<sup>2</sup> | --<sup>2</sup> | 0.67<br>(0.19, 2.43) | 0.70<br>(0.20, 2.50) | 0.57<br>(0.17, 2.07) | 0.61<br>(0.18, 2.12) |
| Model fit statistics |  |  |  |  |  |  |
| Log likelihood | -273.48 | -266.64 | -924.31 | -902.74 | -945.57 | -924.48 |
| Model p-value | 0.033 | 0.028 | <0.001 | <0.001 | <0.001 | <0.001 |
| Akaike's Information Criteria (AIC) | 560.96 | 557.28 | 1864.61 | 1831.48 | 1883.43 | 1874.96 |
| Bayesian Information Criteria (BIC) | 605.31 | 633.18 | 1915.92 | 1914.71 | 1934.66 | 1958.31 |

RDT = rapid diagnostic test; PCR = polymerase chain reaction; OR = odds ratio; CI = confidence interval; MC = malaria camp
<sup>1</sup> Adjusted for age, gender, education, and occupation
<sup>2</sup> No RDT malaria at follow-up 3; ORs small and unstable

### Intent-to-treat analysis for secondary outcomes

The anthropometric and clinical outcomes at baseline and the third follow-up visit are shown in **Table 4**; associations are considered significant if p<0.013. At baseline, 33.3% were severely underweight, with no differences across study arms. At follow-up, the prevalence of severe underweight increased to 36.8%, but this increase was not significant; there were still no differences across arms in underweight at follow-up. Few (2.2%) were severely anemic, with significantly more anemia in Arm A (5.1%) as compared to Arms B and C (0.1 and 0.7%, respectively; p<0.001). At follow-up there were no significant differences in anemia across arms, but there were significant decreases from baseline to follow-up for the total sample and Arm A. At baseline, prevalence of fever was 24.3%. Arm B had the highest prevalence (38.1%) as compared to Arm A (14.6%) and Arm C (13.9%, p<0.001). There were no significant differences in fever across arms at follow-up, although there were significant reductions from baseline to follow-up overall and for each arm (p<0.001). Prevalence of malnutrition measured by MUAC was 19.3% at baseline, with higher prevalence in Arm C (26.4%) as compared to Arms A (18.2%) and B (17.2%, p<0.001). At follow-up there were significant differences in malnutrition across arms but no significant differences from baseline to follow-up. Crude and adjusted multilevel mixed effects logistic regression models were not estimated for these secondary outcomes due to small numbers (i.e., severe anemia and fever) and/or few significant differences in the bivariate analyses comparing baseline to follow-up (i.e., severe underweight, severe anemia, MUAC malnutrition).

**Table 4.** Anthropometric outcomes at baseline and the third follow-up visit

| Variable | Total sample<br>N=2463<br>n (%) | Arm A<br>New MC<br>n=1001<br>n (%) | Arm B<br>Delayed MC<br>n=1023<br>n (%) | Arm C<br>Old MC<br>n=439<br>n (%) | p-value<sup>1</sup> |
| --- | --- | --- | --- | --- | --- |
| **Baseline** |  |  |  |  |  |
| Severe underweight |  |  |  |  | 0.068 |
| No | 1633 (66.7) | 655 (66.3) | 665 (65.1) | 313 (71.3) |  |
| Yes | 815 (33.3) | 333 (33.7) | 356 (34.9) | 126 (28.7) |  |
| Missing | 15 | 13 | 2 | 0 |  |
| Severe anemia |  |  |  |  | <0.001<sup>2</sup> |
| No | 2409 (97.8) | 951 (95.0) | 1022 (99.9) | 436 (99.3) |  |
| Yes | 54 (2.2) | 50 (5.1) | 1 (0.1) | 3 (0.7) |  |
| Missing | 0 | 0 | 0 | 0 |  |
| Fever |  |  |  |  | <0.001 |
| No | 1863 (75.7) | 852 (85.4) | 633 (61.9) | 378 (86.1) |  |
| Yes | 597 (24.3) | 146 (14.6) | 390 (38.1) | 61 (13.9) |  |
| Missing | 3 | 3 | 0 | 0 |  |
| MUAC malnutrition<sup>3</sup> |  |  |  |  | <0.001 |
| No | 1983 (80.7) | 814 (81.8) | 846 (82.8) | 323 (73.6) |  |
| Yes | 473 (19.3) | 181 (18.2) | 176 (17.2) | 116 (26.4) |  |
| Missing | 7 | 6 | 1 | 0 |  |
| **Third follow-up visit** | 2007 (81.5) | 813 (81.2) | 828 (81.0) | 366 (83.4) | 0.541 |
| Severe underweight |  |  |  |  | 0.252 |
| No | 1265 (63.4) | 511 (63.3) | 510 (61.8) | 244 (66.9) |  |
| Yes | 732 (36.7) | 296 (36.7) | 315 (38.2) | 121 (33.2) |  |
| Missing | 10 | 6 | 3 | 1 |  |
| p-value<sup>4</sup> | 0.019 | 0.141 | 0.189 | 0.173 |  |
| Severe anemia |  |  |  |  | 0.590<sup>2</sup> |
| No | 2004 (99.9) | 812 (99.9) | 827 (99.9) | 365 (99.7) |  |
| Yes | 3 (0.2) | 1 (0.1) | 1 (0.1) | 1 (0.3) |  |
| Missing | <0.001<sup>2</sup> | 1.000<sup>2</sup> | <0.001<sup>2</sup> | 0.630<sup>2</sup> |  |
| p-value<sup>2</sup> |  |  |  |  |  |
| Fever |  |  |  |  | 0.225<sup>2</sup> |
| No | 1984 (99.4) | 800 (99.1) | 819 (99.3) | 365 (100.0) |  |
| Yes | 13 (0.7) | 7 (0.9) | 6 (0.7) | 0 (0.0) |  |
| Missing | 10 | 6 | 3 | 1 |  |
| p-value<sup>2</sup> | <0.001 | <0.001 | <0.001 | <0.001<sup>2</sup> |  |
| MUAC malnutrition<sup>3</sup> |  |  |  |  | 0.004 |
| No | 1662 (83.2) | 693 (85.9) | 684 (82.8) | 285 (78.1) |  |
| Yes | 336 (16.8) | 114 (14.1) | 142 (17.2) | 80 (21.9) |  |
| Missing | 9 | 6 | 2 | 1 |  |
| p-value<sup>2</sup> | 0.036 | 0.987 | 0.020 | 0.138 |  |

MC = malaria camp; BMI = body mass index; MUAC = mid-upper arm circumference
<sup>1</sup> $\chi^2$ p-value for comparisons across arms<sup>2</sup> Fisher's exact test<sup>3</sup> <23 cm for adults and adolescents aged 15 and older, <11.5cm for children 6 to 60 months, <12.9cm for 5-9 year olds, <16cm for 10-14 year olds<sup>4</sup> $\chi^2$ p-value for comparing baseline to follow-up 3, unless otherwise noted

### *Pfhrp2* deletion prevalence in study sample

From a total of 2,463 individuals enrolled in our baseline survey across all study arms, 240 (9.7%) were infected with *Plasmodium* as detected by RDT and/or PCR at baseline, and 192 (80%) of those were RDT-/PCR+ (*i*.*e*., subpatent). Because the prevalence of subpatent infections was so high, we undertook assays to determine whether there were deletions of the *pfhrp2* gene among the cases. *Pfhrp2* deletions are a major contributor to RDT failures both globally^11^ and in the Odisha region^12, 13, 14^, since most of the commercially available RDTs detect histidine-rich protein 2 (HRP2) which is produced during the asexual blood stage of *P. falciparum* infection.

Of the n=113 fever positive RDT-/PCR+ infections from the baseline survey, 95 (84.1%) were *pfhrp2* negative, 2 (1.8%) were *pfhrp2* positive, and 13 (11.5%) were inconclusive (**Table 5**). An additional 79 *P. falciparum* RDT-/PCR+ fever negative baseline infections were tested: 41 (51.9%) were *pfhrp2* negative and 32 (40.5%) were *pfhrp2* positive. A total of n=62 RDT- /PCR+ infections were identified across the FU1, FU2, and FU3 surveys, of which 19 (30.7%) were *pfhrp2* negative and 40 (64.5%) were *pfhrp2* positive.

**Table 5.** *pfhrp2* gene deletion prevalence in subpatent (RDT-/PCR+) infections.

|  | Baseline<br>Total | Fever | No fever | Any follow-up |
| --- | --- | --- | --- | --- |
| Number of infections | 192 | 113 | 79 | 62 |
| *Pfhrp2* negative | 136 (70.8) | 95 (84.1) | 41 (51.9) | 19 (30.7) |
| *Pfhrp2* positive | 34 (17.7) | 2 (1.8) | 32 (40.5) | 40 (64.5) |
| Inconclusive | 13 (6.8) | 13 (11.5) | 0 (0.0) | 0 (0.0) |
| Not tested | 9 (4.7) | 3 (2.7) | 6 (7.6) | 3 (4.8) |

RDT = rapid diagnostic test; PCR = polymerase chain reaction

### Malaria exposure and serology in a subset of study participants

We assayed 300 baseline samples (240 PCR+ and 60 PCR-) for anti-*Plasmodium* antibodies to 13 *P. falciparum*-specific antigens and four *P. vivax*-specific antigens using the Luminex MAGPIX platform and xPONENT software to estimate mean fluorescence intensity (MFI). Data were missing for six markers (n=180 missing for PvMSP10; n=20 missing for PfEBA140 R3-5, PfETRAMP5_ag1, PfMSP2CH150, PfRh2_2030, and PvMSP8). There were no significant differences by gender for *P. falciparum* and *P. vivax* antibodies, but the proportion positive increased as age increased (data not shown).

**Table 6** summarizes the antibody results by study arm and infection status. There were no significant differences between Arms A and B in terms of *P. falciparum* and/or *P. vivax* antibodies at baseline. There were significant differences in antibodies across all three arms, with a lower proportion of Arm C participants having *Plasmodium* antibodies. With respect to baseline infection status, a higher proportion of symptomatic participants had *P. falciparum* antibodies as compared to uninfected and asymptomatic participants. A higher proportion of asymptomatic participants had *P. vivax* antibodies as compared to uninfected and infected participants.

**Table 6.** Baseline Plasmodium antibodies among 240 PCR+ and 60 PCR-participants in the Phase 1 study, by study arm and infection status

|  | Any Pf antibodies<br>(n=189) |  | Any Pv antibodies<br>(n=183) |  | Any Pf or Pv antibodies<br>(n=191) |  |
| --- | --- | --- | --- | --- | --- | --- |
|  | n (%) | p | n (%) | p | n (%) | p |
| Study arm |  |  |  |  |  |  |
| Arm A (New MC) | 109 (62.6) | 0.022* | 46 (26.4) | 0.001* | 111 (63.8) | 0.021* |
| Arm B (Delayed MC) | 69 (69.7) | 0.240** | 36 (36.4) | 0.085** | 69 (69.7) | 0.322** |
| Arm C (Old MC) | 11 (40.7) |  | 1 (3.7) |  | 11 (40.7) |  |
| Infection status |  | <0.001 |  | 0.011 |  | <0.011 |
| Uninfected | 26 (55.3) |  | 13 (27.7) |  | 26 (55.3) |  |
| Symptomatic | 91 (77.8) |  | 43 (36.8) |  | 91 (77.8) |  |
| Asymptomatic | 26 (55.3) |  | 91 (77.8) |  | 74 (54.4) |  |

\* Arm A vs. B vs. C \*\* Arm A vs. B PCR = polymerase chain reaction; Pf = *Plasmodium falciparum*; Pv = *Plasmodium vivax*; MC = malaria camp

### Village-level costs of MCs

We conducted an analysis of the implementation costs of MCs in the study villages from the service provider perspective. The results of the cost analysis are presented in **Table 7**.

In the study villages, the cost per person ranged between US$3-8, the cost per tested between US$4-7, and the cost per treated between US$82-1,614 per camp round.

**Table 7.**
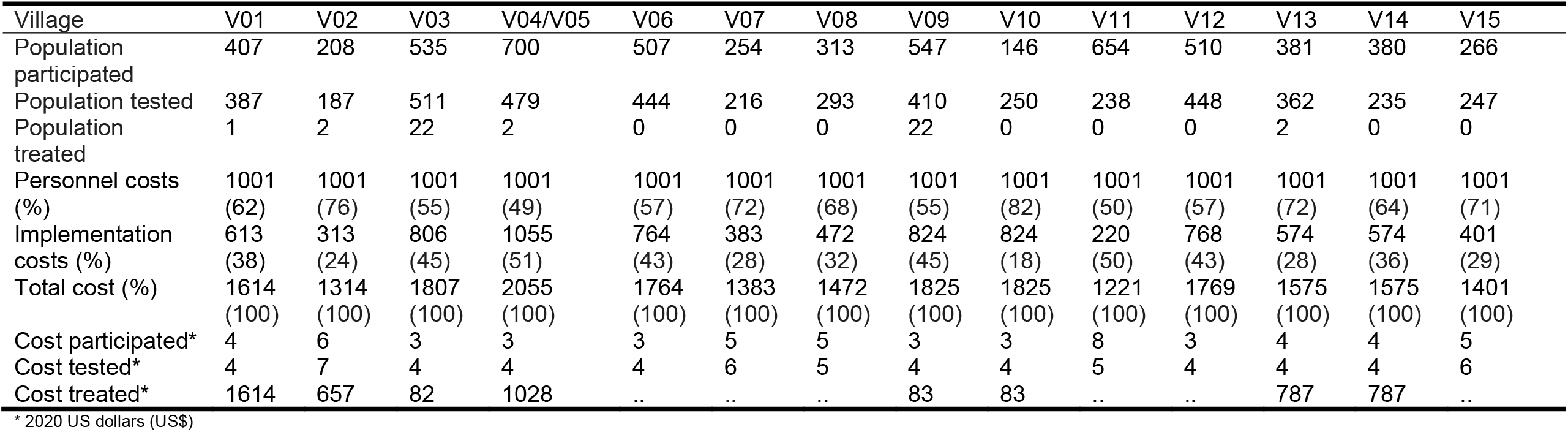
Village-level costs of the DAMAN intervention in study villages in Odisha State, India (all costs are in 2020 US dollars; *per person)

## Discussion

In this quasi-experimental study conducted in rural villages in Odisha, intent-to-treat analyses suggest that the DAMaN program’s malaria camps were associated with lower malaria incidence relative to standard-of-care. This is suggested by the reductions in malaria infection prevalence at baseline versus FU3. We note that all villages received MCs at some point, with Arm A receiving MCs four times, Arm B receiving it twice, and Arm C receiving it prior to the study’s inception and three times during the study. Intervention activities were impacted by the COVID-19 pandemic-related shutdowns and, as a result, 9 villages (2 of 6 in Arm A, 6 of 6 in Arm B, and 2 of 3 in Arm C) did not receive any malaria camp activities during the first follow-up. There were no serious adverse events.

Our findings are in contrast to a recent meta-analysis that noted a marginal and non-significant pooled effect size for MSAT interventions on malaria infection prevalence and incidence.^15^ One major limitation of previous evaluations of MSAT is low sensitivity of the diagnostic methods (i.e., RDTs and light microscopy)^15^. The Odisha DAMaN MCs use RDTs for MSAT while this evaluation used both an RDT and PCR, overcoming some previous evaluations’ limitations.

There were no significant differences in the subset of Arm A and B participants tested for *Plasmodium* antibodies, suggesting that Arms A and B had similar malaria exposure prior to the intervention. There were significant differences in *Plasmodium* antibodies across all 3 arms, with a lower proportion of Arm C participants having antibodies; this suggests lower transmission of *Plasmodium* in these villages and/or impact of the previous exposure to the MC intervention.

At baseline, more febrile participants had *P. falciparum* antibodies as compared to uninfected and asymptomatic participants, while more afebrile participants had *P. vivax* antibodies. These results suggest possible cross-reactivity of *P. falciparum* and *P. vivax* antibodies and antigens in the assay as well as potential previous exposure to *P. vivax*. The lower proportions of participants with *P. vivax* antibodies relative to *P. falciparum* antibodies is consistent with RDT and PCR data demonstrating lower prevalence of *P. vivax* in this study sample, as well as data from the National Center for Vector Borne Diseases Control showing that 91.4% of malaria cases in 2021 were *P. falciparum* infections^3^.

The Malaria Control Program implemented MSAT across all malaria camp rounds rather than FSAT in the subsequent rounds. This likely contributed to the impact of the intervention, as 54 of the 74 RDT+ malaria cases (73.0%) detected in the study were afebrile [data not shown]. In other words, they treated more malaria cases than they otherwise would have if they had only tested and treated febrile cases each time.

With regard to the distribution of LLIN as part of the malaria camps, a separate, small survey of 151 households undertaken in the 15 study villages found that 93.4% of surveyed households already had at least one bed net, of which 97.9% reported that it was treated with insecticide [data not shown]. So, although LLINs were not distributed during these malaria camp rounds and we did not collect data on LLIN or general net ownership or use in the survey, it is possible that many households already had them.

The most common commercial RDTs in India target the Pfhrp2/3 antigens. We observed a high prevalence of the *pfhrp2* gene deletion, and although this had been previously reported in Odisha in 2014 it was not thought to be widespread at the time^12^. A cross-sectional study using samples from 2013-2016 found parasites with deletions throughout Odisha state^14^ including the Northern Plateau where the study sites are located. Pati *et al*.^14^ reported a 15% RDT failure rate in that study; of the 58 RDT-/PCR+ samples, 65.5% were *pfhrp2* negative. Bharti *et al*. ^12^ reported that 66.7% of RDT-/PCR+ samples were *pfhrp2* negative in the Koraput district of Odisha in 2014. In our study, approximately 80% of PCR+ *Plasmodium* infections were not detected by the RDT and ∼71% of RDT-/PCR+ *P. falciparum* infections were *pfhrp2* negative.

The World Health Organization recommends implementing RDTs targeting the PfLDH antigen in settings with a high prevalence of *pfhrp2/3* gene deletions^16^; findings from this and previous studies suggest that alternative diagnostic approaches are now needed in this region as interventions that rely solely on *pfhrp2* RDTs for the detection of infections may miss the majority of *P. falciparum* infections which predominate in Odisha.

The cost per person for the MCs ranged between US$3-8, the cost per tested person between US$4-7, and the cost per treated person between US$82-1,614 per MC round.

According to our systematic literature review on the effectiveness and cost-effectiveness of intermittent mass MSAT interventions for malaria^17^, these results are comparable to the costs reported in the published literature, ranging between US$3.5-14.3 per person tested per round, and corresponds roughly to 20-35% of the per-capita domestic general health expenditure in India (US$19.6 in 2018)^18^.

We note several important study limitations. First, as discussed in our study protocol^7^, the villages were not randomized to study arms, which may result in potential selection bias. Second, COVID-19 pandemic shutdowns resulted in disruption in delivery of the MCs at FU1 which meant that several villages may not have received all or any of the activities in that MC round. Third, while the follow-up was over 80%, people with *Plasmodium* infection at baseline were less likely to be followed-up but there were no significant differences in follow-up among asymptomatic or subpatent cases.

Collectively, our quasi-experimental cluster-assigned stepped-wedge trial results suggest that the DAMaN malaria camp intervention is associated with reductions in malaria in rural villages and thus is a promising, financially feasible approach for malaria control in rural settings. However, given the quasi-experimental design, COVID-related study challenges, and *pfhrp2* deletion prevalence, we cannot infer causality. A large-scale cluster-randomized trial of an adapted malaria camp intervention that will address the diagnostic limitations of the current intervention (*i*.*e*., reliance upon RDTs that detect the Pfhrp2/3 antigens) is being planned. This is timely considering that Odisha State government extended the DAMaN initiative in October 2022 for five more years in 21 districts in a bid to achieve malaria elimination in Odisha by 2030^19^.

## Methods

### Ethics statement

Ethical approval for the trial was obtained from the Odisha State Research and Ethics Committee (Odisha, India), institutional ethics committee at Community Welfare Society Hospital (Rourkela, Odisha, India) and the institutional review board at New York University (New York, NY, USA). All research was performed in accordance with relevant guidelines/regulations and the Declaration of Helsinki.

### Study design and enrollment procedures

The quasi-experimental cluster-assigned stepped-wedge study design, power analysis, and recruitment/enrollment procedures are described in a protocol paper^7^ and the study was registered at ClinicalTrials.gov (NCT03963869). Briefly, 12 intervention villages were selected in two districts of Odisha state, Keonjhar and Jharsuguda, and distributed in equal numbers between two study arms: Arm A ‘new-MC’ (communities receiving MCs for the first time in year one) and Arm B ‘delayed-MC’ (communities undergoing routine malaria control in year one and receiving MCs for the first time in year two); control Arm C contained three ‘old-MC’ villages, where MCs had already been implemented prior to the study period. A cohort was planned with target enrollment of 2,700 individuals across all 15 villages (i.e., 1100 in Arms A and B; 500 in Arm C).

Study participants in each village were recruited and enrolled at baseline before the initial administration of the intervention and subsequently surveyed prior to each round of screen and treat by the Government of Odisha DAMaN team. Study participants in the three villages not receiving the intervention were recruited and enrolled in parallel temporally. Residents of the study villages aged 1 to 69 were eligible for the study. Written informed consent was obtained in the local language from all adult study participants aged 18-69 years with apparent full comprehension of the study procedures. Assent was obtained in addition to parental or legal guardian informed consent for participants aged 7-17 years. Parental informed consent was obtained for children aged 1-6 years. During each survey round, all subjects completed a health questionnaire that captured demographic, malaria knowledge and prevention, and malaria infection and treatment history data, and provided a blood sample for *Plasmodium* parasite detection.

### Intervention adaptations due to COVID-19 pandemic

Arm A (new MC) and C (old MC) villages were to receive two cycles of MCs, each with two rounds of MSAT. The first MC cycle was from August to November 2019 (Round 1) and January to March 2020 (Round 2). We note that the Round 2 MSAT was interrupted by COVID-19 pandemic shutdowns. The second MC cycle was from June to September 2020 (Round 1) and October to December 2020 (Round 2). We note that Round 1 MSAT was also interrupted by COVID-19 pandemic shutdowns. Arm B (delayed MC) villages were to receive SOC in the first cycle (August 2019 – March 2020) and one MC cycle with two rounds of MSAT from June to September 2020 (Round 1) and October to December 2020 (Round 2). Again, Round 1 was interrupted by the pandemic.

### Primary outcomes

The primary outcome measures were any *Plasmodium* as detected by PCR and *Plasmodium* species (i.e., *P. falciparum* and/or *Plasmodium vivax*) as detected by PCR. Secondary malaria outcome measures included *Plasmodium* infection as detected by RDT and *Plasmodium*-specific serology. *Plasmodium* infection by RDT is defined as RDT negative (RDT-) or RDT positive (RDT+). We further classified infections as ‘asymptomatic’, defined as PCR+ and/or RDT+ for any *Plasmodium* species with absence of documented fever or self-reported fever in the last 48 hours, and/or ‘subpatent’, defined as RDT- and PCR+. *Plasmodium*-specific serology was analyzed as a continuous antibody titer variable as well as a categorical (seropositive vs. seronegative) variable.

### Blood sample collection and processing

Blood samples for blood smears, blood spots (Whatman FTA cards), a small volume microvette, and micro volumes required for point of care hemoglobin and RDTs, were collected from consenting study participants by sterile lancet finger prick as previously described^7^. Vacutainers (5 ml; BD Vacutainer glass blood collection tubes with acid citrate dextrose) of venous blood were requested from study participants found to be positive by RDT. Microvette and vacutainer samples were refrigerated until separated into plasma and infected red blood cell (iRBC) components by centrifugation. Plasma was stored at - 80°C until assayed for *Plasmodium* immunoglobulin G (IgG), and total DNA was extracted from the iRBC pellet using QIamp DNA mini kit (QIAGEN) with a final elution volume of 50 μl.

### Molecular detection of *Plasmodium* parasites by species-specific PCR

DNA samples were tested for *Plasmodium* species by a PCR assay targeting multi-copy loci Pfr364 and Pvr47^20^ as previously described^7^. Five μl of DNA was used in a total reaction volume of 30 μl, and amplification products were visually confirmed by ethidium bromide-stained gel electrophoresis using a Gel Doc EZ documentation system (BioRad Laboratories, Inc.).

### Detection of anti-*Plasmodium* IgG by Luminex MAGPIX cytometric bead array

Plasma isolated from a subset of microvette samples of finger prick blood was assayed for 13 anti-*P. falciparum* and four anti-*P. vivax* antibodies as previously described.^7^ The panel included antigens whose corresponding antibodies have been suggested as indicators of protection from clinical disease^21, 22^, long-term or cumulative exposure^23, 24, 25, 26^, and recent infection^23, 24, 25, 26^. Briefly, n=300 plasma samples were diluted 1/200 and assayed in duplicate according to standard procedures^27, 28^. xPONENT software was used for data acquisition (Luminex Corp., Austin, TX). Specifically, the net Mean Fluorescence Intensity (MFI; net MFI_Ag_ = raw MFI_Ag_ - background MFI_Ag_ where background MFI_Ag_ is the mean MFI of a given antigen in the blank wells) was calculated for each antigen in each sample. The presence of antibody to each antigen (e.g., seropositivity) was defined as the mean net MFI_naive pool_ plus three standard deviations. Composite variables for any *P. falciparum*, any *P. vivax*, and any *P. falciparum* or *P. vivax* antibodies were created.

### Secondary anthropometric and clinical outcomes

We report on several anthropometric measurements, including severe underweight, severe anemia, fever, and malnutrition as measured by mid-upper arm circumference (MUAC). For adults aged 18 and older, severe underweight was defined as a BMI<16. For children, severe underweight was defined as three standard deviations below the median based on the WHO growth charts used for children <5 years and the revised Indian Academy of Pediatrics (IAP) growth charts for Indian children and adolescents aged 5 to 17^29^. Severe anemia was defined as hemoglobin <5 g/dl for persons aged <12 and 7 g/dl among those aged 12 and older^30^. Fever was defined as a temperature of 99.5°F or higher. MUAC malnutrition was defined as <23 cm for adults and adolescents aged 15 and older^31, 32^, <16 cm for 10-14 year olds^33^, <12.9 cm for 5-9 year olds^33^, and <11.5 cm for children 6 to 60 months^34^.

### Identification of *pfhrp2* gene deletions

DNA extracted from microvette samples from RDT- /PCR+ individuals across all survey rounds were evaluated for *pfhrp2* gene deletions using a multi-step workflow modified from Bharti *et al*., and Baker *et al*.,^12, 13^. Briefly, the workflow consisted of (1) conventional PCR targeting *pfhrp2* exon 2 of all RDT-/PCR+ samples, (2) nested PCR targeting *pfhrp2* exon 1-2 of all *pfhrp2* exon 2 positive samples, (3) nested PCRs targeting the upstream and downstream flanking regions of all *pfhrp2* exon 1-2 positive samples, and (4) a DNA sample quality control confirmation qPCR of *pfhrp2* exon 2, exon 1-2, and/or flanking region negative samples using screening of the Pf *aldolase* gene. A total of n=271 RDT-/PCR+ DNA samples were evaluated.

### Costing study design, data collection, and analyses

Using an activity-based costing method, we conducted an analysis of the implementation costs of malaria camps (MCs) in the study villages from the service provider perspective.^35^ Interviews were conducted with the malaria camp supervisors who led the MCs in the study villages, and programmatic documentation from the Odisha State malaria control program were reviewed to identify the activities related to the planning and implementation of DAMaN at the village-level. These included: (1) Training activities (training of public health supervisors and workers as well as village volunteers, including the village specific ASHAs; (2) Micro-plan development for MCs; (3) Planning and sensitization activities (identification of village volunteers, selection of camp venue and time); (4) Community sensitization activities; (5) Community mobilization activities; (6) Health activities (malaria testing and treatment, child health, maternal health); (7) Monitoring and supervision activities; and (8) Follow-up activities. Resource use was linked to specific activities associated with an intervention. During interviews, the MC supervisors identified the time dedicated by MC staff to different activities for each camp round. Time costs of MC staff were calculated based on average annual gross wage rates, apportioned according to time devoted to DAMaN activities per camp round, and summed across all MC staff. Unit costs of rapid diagnostic kits and antimalarial drugs were extracted from the DAMaN Operational Guidelines^36^. The costs of these commodities were calculated by multiplying the quantities used during a camp round by their unit costs based on participation in each camp round in a given village. Personnel and implementation costs per round were summed and divided by the population that participated in the MC, the population tested for malaria, and the population treated for malaria, in each study village to calculate cost per person, cost per person tested, and cost per person treated per camp round. All costs were collected in local currency and converted to and presented in 2020 US dollars (US$), using the average exchange rate for 2020 (1 US$ = 74.27 Indian Rupees)^37^.

### Statistical analyses

The primary outcome was *Plasmodium* presence and species as detected by PCR at the third follow-up visit. Secondary outcomes included *Plasmodium* presence and species as detected by RDT and anthropometric and clinical outcomes including underweight, anemia, fever, and malnutrition as measured by mid-upper arm circumference (MUAC) at the third follow-up visit. Missing data was minimal among the analytic sample and ranged from 0 to 1.5% depending on the variable.

Bivariate analyses compared outcomes and key covariates across study arms with Pearson’s χ^2^ or Fishers exact tests (if cell sizes were <5) at baseline and outcomes at follow-up 3. We use a Bonferroni adjusted α of 0.013 to account for multiple comparisons in the bivariate analyses comparing baseline to follow-up outcomes. Follow-up rates were calculated as the proportion of baseline participants that were followed up at the third time point.

Multilevel mixed-effects logistic regression was used to estimate the intervention effects, which enabled us to account for the repeated measure and nesting of individuals within the 15 villages. An interaction term (study arm x visit [baseline vs. FU3]) was used to estimate time effects for each arm and village was included as a random effect. We first estimated the crude associations for each outcome by study arm. We then adjusted these estimates for age, sex, education, and occupation. Odds ratios with 95% confidence intervals are reported. All analyses were conducted with Stata 17.0 (StataCorp, College Station, TX, USA) and were intent-to-treat. The study was registered at ClinicalTrials.gov (#NCT03963869).

## Data availability

Epidemiology study data are available in the Clinical Epidemiology Database^38^ (https://clinepidb.org) under the ‘India ICEMR DAMaN Quasi-experimental Stepped-wedge’ study title.

## Data Availability

Epidemiology study data are available in the Clinical Epidemiology Database (https://clinepidb.org) under the India ICEMR DAMaN Quasi-experimental Stepped-wedge study title.

## Acknowledgements

We thank study participants and the DAMaN teams in the study villages of Keonjhar and Jharsuguda districts for their cooperation, and Dr Deben Das, Mr. Baladev Nanda, and Mr. Birat R Padhan of the Odisha State Malaria Control Programme for their support. We thank field team members Asit Samal, Anna Bage, Pratibha Ekka, Satyanarayana Panigrahi, Sahil Ekka, John P Lakra, Sobharani Toppo, and Madhusmita Parichha and lab technicians Swagatika Dash and Satyaranjan Chhatria at Community Welfare Society Hospital for their efforts, and Dr. Kevin Tetteh, Ms. Elin Dumont, and Ms. Catriona Patterson of the London School of Hygiene and Tropical Medicine (England, UK) for providing serology assay support. We also thank Dr. James Beeson (Burnet Institute, Australia) and Dr. Simon Draper (University of Oxford, UK) for providing serology assay antigens. This study was supported by the National Institute of Allergy and Infectious Diseases (NIAID) of the National Institutes of Health (NIH) under Award Number U19AI089676 as part of the International Centers of Excellence for Malaria Research. The content does not necessarily represent the official views of the US NIH or NIAID.

## Author contributions

DO, AK, AVE, YT, SAS, PKS, SM1 (Sanjib Mohanty), MMP, and JMC conceptualized and developed the study design and methodology with input from all authors. PKS led the field and laboratory work in Odisha, India with support from TKP, MAH, SM, and MMP. SM2 (Stuti Mohanty) and PKS completed the *pfhrp2* gene deletion assay with support from AK. AK undertook the serology assay. DO and SAS managed the data. DO, AJ, and YT completed the data analyses. JMC acquired the funding. DO, AK, YT, JMC, and PKS wrote the manuscript with input from all authors.

## Competing interests

Dr. Madan M. Pradhan is employed by the Odisha State Vector Borne Disease Control Programme, which is the organization that is implementing the intervention. He has a potentially self-serving stake in the research results via potential promotion or career advancement based on outcomes. The other authors have no competing interests to declare.

## Notes

### Clinical Trial

NCT03963869

### Clinical Protocols

https://pubmed.ncbi.nlm.nih.gov/33866961/

